# Effects of a High-Airflow Fan-Attached Garment on Thermoregulatory and Physiological Responses During Physical Activity in Hot Environments: A Randomized Crossover Trial

**DOI:** 10.64898/2026.11.23.26368461

**Authors:** Kazuma Hayashi, Megumi Wanifuchi, Hironaga Ito, Yuki Urushizawa, Yoshihito Tomioka, Fuminori Takayama

## Abstract

**Objectives:** Although fan-attached garments are widely used, conventional airflow rates have shown limited efficacy in suppressing core body temperature elevation during physical activity. We aimed to investigate the effects of a high-airflow fan-attached garment on core body temperature and physiological strain during physical activity in hot environments under different humidity levels.

**Methods:** Seven healthy adult men completed four randomized crossover trials involving two environmental conditions—wet bulb globe temperatures of 25°C (36°C, 20% relative humidity) and 31°C (36°C, 60% relative humidity)—and two garment conditions: control and fan-attached garment (80 L/s airflow). Each trial consisted of a 10-min thermoneutral rest, 10-min hot rest, and 60-min continuous cycle ergometer exercise (60 W). We evaluated rectal and skin temperatures, heart rate, microclimate environment, body mass change, and subjective sensations.

**Results:** At a wet bulb globe temperature of 31°C, the fan-attached garment significantly suppressed the increase in rectal temperature compared with the control garment at 60 min of exercise (difference: 0.50°C, p < 0.001), with rectal temperature remaining unchanged from baseline (+0.11°C, p = 0.077). Exercise heart rate (difference: 31.5 beats/min, p < 0.001) and body mass loss (difference: 0.5 kg, p = 0.001) were also significantly reduced with the fan-attached garment. Microclimate humidity was significantly lower with the fan-attached garment throughout exercise in both environments (p < 0.001).

**Conclusions:** The high-airflow (80 L/s) fan-attached garment effectively prevented an increase in core body temperature and mitigated cardiovascular and thermoregulatory strain during moderate physical activity, even in hot and humid environments.

**Trial registration number:** UMIN000061936

*Key points:* 

*What is already known on this topic:* Fan-attached garments are widely used to reduce heat strain; however, evidence regarding the efficacy of high-airflow fans (e.g., 80 L/s) remains limited.

*What this study adds:* Wearing a fan-attached garment equipped with a high-airflow fan (80 L/s) substantially lowered clothing microclimate humidity under low humidity and suppressed its increase under high humidity, and almost completely prevented an increase in core body temperature (+0.11°C) during 1 h of moderate exercise in a hot and humid environment (wet bulb globe temperature 31°C; 36°C, 60% relative humidity). Furthermore, it markedly reduced heart rate (by up to 32 bpm) and body mass loss (by 0.5 kg).

*How this study might affect research, practice or policy:* High-airflow fan-attached garments provide an effective heat-mitigation strategy for industrial workers performing continuous physical labor in hot environments. These findings support practical workplace interventions to safeguard workers’ health and sustain productivity in accordance with regulatory heat-prevention guidelines.

## 1. Introduction

As global warming and climate change progress, occupational labor in hot environments is a major challenge for health management and productivity maintenance^1^. Excessive increases in core body temperature induce direct cytotoxicity and coagulopathy, potentially progressing to severe multiple organ dysfunction and even death^2^. Physical work performed in hot conditions without adequate mitigation can decrease productivity^3^. Thus, effective thermoregulatory and heat-stress mitigation strategies are urgently required.

Personal cooling garments (PCGs) mitigate heat stress and are categorized into conductive, evaporative, convective, radiative, and hybrid systems^4^. Among these, fan-attached garments, classified as convective cooling systems that force ambient air into the garment using small electric fans, are widely used by industrial workers. They mitigate increases in core body temperature and heart rate^5,6^. However, at conventional airflow rates, their capacity to suppress increases in core body temperature during physical activity in hot environments is limited^5,6^. The heat dissipation capacity does not fully compensate for metabolic heat production during physical activity, thereby failing to prevent excessive core body temperature increases^5,6^. Evaporative cooling under clothing requires a reduction in microclimate humidity to provide a meaningful enhancement of heat dissipation capacity. In a previous study, when the difference in microclimate humidity was approximately 10%, no substantial effect on systemic physiological responses, such as core body and skin temperatures, was observed^7^. Thus, a marked reduction in microclimate humidity is essential for meaningful thermoregulation^8^.

In high-temperature environments equal to or exceeding skin temperature (approximately ≥35°C), fan airflow may draw in hot air, increasing convective heat gain and elevating core temperature^9^. Evaluating the effects of fan-attached garments at high temperatures exceeding 35°C, where such convective heat gain may occur, is critical for efficacy and occupational safety. In hot environments, human heat dissipation relies on sweat evaporation. Although fan airflow can enhance evaporative heat loss even under high humidity during rest while wearing light clothing^10^, when work garments are worn or physical activity is performed, high ambient humidity may reduce the drying capacity of the drawn-in air; consequently, sweat evaporation may be impaired, leading to a plateau in fan cooling efficiency.

Non-fan-attached PCGs include passive garments that rely on the sweat-absorbing, quick-drying, and breathable properties of functional fabrics^8^, and conductive cooling modalities such as water-circulating cooling vests^6^ and phase change materials (PCMs)^11^; however, evidence that they consistently suppress core body temperature elevation is limited. Moreover, they present ergonomic and operational challenges in occupational settings, including increased garment weight, restricted mobility, and limited cooling duration.

Garments equipped with high-airflow fans (e.g., 80 L/s), which exceed conventional airflow capacities, are commercially available. This technology expels humid air from the microclimate, resulting in a more pronounced reduction in microclimate humidity than with conventional garments, which may suppress increases in core body temperature. However, no studies have rigorously examined the physiological effects of high airflow on core body temperature (rectal temperature) and microclimate in a controlled hot environment.

We examined the effects of a high-airflow fan-attached garment on core body temperature during 1 h of continuous physical activity in healthy adult men in a hot environment (36°C) across two different humidity conditions: low humidity (20% relative humidity [RH], corresponding to wet bulb globe temperature [WBGT] 25°C) and high humidity (60% RH, corresponding to WBGT 31°C).

## 2. Methods

### 2.1 Participants

In this randomized crossover trial, we included healthy adult men aged ≥18 and <46 years who provided written informed consent, had a body mass index of 18.5–25 kg/m^2^, regular exercise habits of at least twice weekly, and the ability to understand the study procedures and comply with all requirements.

We excluded individuals with current treatment for any illness; a history of serious diseases requiring pharmacotherapy, physician-diagnosed heat-related illness (Grade II or higher), physician-diagnosed vasovagal reflex, or syncope following exercise; orthopedic complaints or prior trauma that could interfere with cycle ergometer exercise; routine use of medications or dietary supplements affecting thermoregulation, circulatory function, or metabolism; current smoking habit; skin conditions that interfere with the attachment of measurement devices or allergies to the test materials; and those judged unsuitable by the principal investigator or study physician.

Of the eight men who provided consent, one was excluded for not meeting the inclusion criteria; thus, seven men (age: 37.4 ± 8.3 years, height: 174.9 ± 4.2 cm, body mass: 72.5 ± 8.1 kg) were included in the final analysis (Supplementary Figure S1). No participant met the trial termination criteria, and no adverse events occurred. This study was approved by the Ethics Committee of Shiba Palace Clinic (approval number: 163084_rn-43266) and conducted in accordance with the ethical standards of the Declaration of Helsinki.

### 2.2 Experimental Design

Each participant completed four experimental trials combining two environmental conditions: WBGT 25°C (ambient temperature: 36°C, relative humidity: 20%) and WBGT 31°C (ambient temperature: 36°C, relative humidity: 60%), and two garment conditions (control [CON] and fan-attached garment [FAN]). The four trials were WBGT 25°C-CON, WBGT 31°C-CON, WBGT 25°C-FAN, and WBGT 31°C-FAN.

In each trial, participants wore standardized base layers consisting of a short-sleeved T-shirt (65% polyester and 35% nylon; UNIQLO CO., LTD., Japan) and long trousers (80% nylon and 20% polyurethane; BURTLE CO., Ltd., Japan). In the CON trials, participants wore a long-sleeved work garment (WA11911; 65% polyester and 35% cotton; SUN-S CO., LTD., Japan). In the FAN trials, participants wore a fan-compatible long-sleeved work garment (KF92651; 65% polyester and 35% cotton; SUN-S CO., LTD., Japan) equipped with a lithium-ion battery (RD9690PJ Japan Label 24V, weight: 345 g; SUN-S CO., LTD., Japan) and two flat fan units mounted on the lower back (RD9620PH 24V, weight: 105 g each; SUN-S CO., LTD., Japan). The fan airflow rate was set at 80 L/s (continuous operating time: approximately 4 h).

The trial order was randomized for each participant. Each experimental session lasted approximately 80 min (Figure 1). The thermoneutral environment (25°C, 50% relative humidity) was established in a constant temperature and humidity chamber (dew-point control system, operating range: 20–25°C, relative humidity: 40–70%; ITECS Co., Ltd., Japan), while the hot environment was maintained in an environmental chamber (dual-coil system, operating range: 5–45°C, relative humidity: 10–85%; ITECS Co., Ltd., Japan).

**Figure 1.**
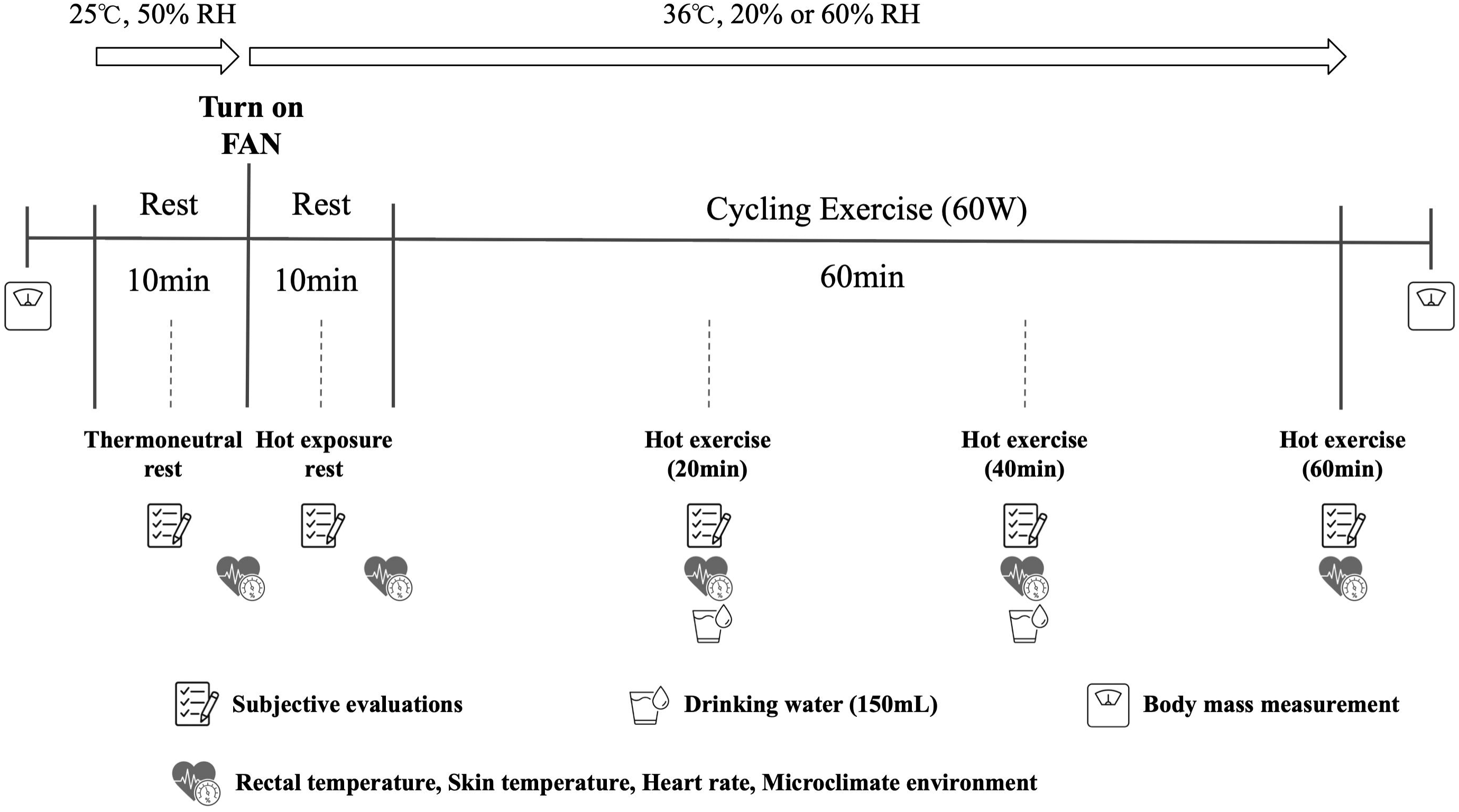
Schematic representation of the experimental protocol. FAN, fan-attached garment; RH, relative humidity.

In the FAN trials, the fans were turned on after the participants completed the seated rest in the thermoneutral environment. The exercise intensity on the cycle ergometer (Fujin-Raijin; OCL Co., Ltd., Japan) was fixed at 60 W with a pedaling cadence of 60 rpm for 60 min. Participants consumed 150 mL of water at 20 and 40 min after the onset of exercise.

Predetermined termination criteria were established to ensure participant safety: 1) core body temperature reaching 39.0°C and persisting for 5 min; 2) participant-reported symptoms of heat-related illness, such as nausea or dizziness; 3) participant request to stop due to volitional exhaustion; or 4) investigator judgment that continuing exercise was unsafe, even in the absence of self-reported symptoms. Participants were instructed to maintain their daily lifestyle habits, abstain from alcohol from the day before each trial, ensure adequate sleep, consume their last meal at least 2 h before testing (consuming only water thereafter), avoid alcohol- and caffeine-containing beverages, refrain from physical exercise for 4 h prior to testing, and avoid exhausting physical activity until the completion of all measurements.

### 2.3 Measurements

Rectal temperature was used as an indicator of core body temperature and served as the primary outcome. A temperature probe (LT-ST08-21 or LT-ST08-31; JP Thermetrics Inc., Japan) was inserted into the rectum and connected to a digital thermometer (LT-8A; JP Thermetrics Inc., Japan) to collect rectal temperature data, which were recorded on a computer using dedicated software (LT-USB1N; JP Thermetrics Inc., Japan). Rectal temperature was recorded at 1-min intervals from the start of seated rest in the thermoneutral environment until exercise completion. Mean values were calculated for 5–10 min of thermoneutral rest, 5–10 min of hot rest, and 15–20, 35–40, and 55–60 min of exercise. Skin temperature, heart rate, microclimate temperature, and microclimate humidity were evaluated at identical time points.

Skin temperature probes (LT-ST08-32; JP Thermetrics Inc., Japan) were attached to the skin surface of the chest and upper arm (directly exposed to fan airflow) and the thigh (not exposed to fan airflow) to measure local skin temperatures. The same thermometer and software were used as for rectal temperature. Probe placement sites were determined based on previous studies^12,13^.

Heart rate was measured using a chest-strap monitor (Polar H10; Polar Electro, Finland) and an application (Polar Beat; Polar Electro, Finland). Microclimate temperature and relative humidity were measured using a temperature/humidity sensor (UNI-01-C003; SANWA SUPPLY INC., Japan) and an application (Wireless temperature and humidity monitor; Step One Limited, Japan). Due to device malfunction, data from one participant in the WBGT 25°C-CON trial and the mean value during the 55–60 min interval from another in the WBGT 31°C-CON trial were missing. Body mass was measured before thermoneutral seated rest and immediately after exercise cessation using a body composition analyzer (BC-768-WH; TANITA Corporation, Japan). The change in body mass was calculated by subtracting post-exercise body mass from pre-test body mass and was used as an indicator of sweat loss.

Subjective outcomes were assessed during thermoneutral rest, hot rest, and at 20, 40, and 60 min of exercise. Thermal sensation was assessed on a 9-point scale (1 = very cold to 9 = very hot)^14^, thermal comfort on a 7-point scale (−3 = very uncomfortable to +3 = very comfortable), rating of perceived exertion (RPE) on the 15-point Borg scale (6 = very, very light to 20 = very, very hard)^15^, and thirst sensation on a 10-point scale (1 = not thirsty at all to 10 = worst imaginable thirst).

### 2.4 Statistical Analysis

Continuous variables were analyzed using linear mixed-effects models and hypothesis testing based on estimated marginal means and 95% confidence intervals (CIs). For core body temperature, skin temperature, heart rate, microclimate temperature, and microclimate humidity, the models included trial condition, time, trial-by-time interaction, trial sequence, and baseline values during thermoneutral rest as fixed effects, with participant ID as a random effect. For body mass change, the model included trial condition, trial sequence, and pre-test body mass as fixed effects. Ordinal variables were analyzed using aligned rank transform (ART) ANOVA to evaluate trial-by-time interactions, followed by Wilcoxon signed-rank tests.

Between-trial comparisons at each time point were conducted for WBGT 31°C-CON versus WBGT 31°C-FAN, WBGT 25°C-CON versus WBGT 25°C-FAN, and WBGT 25°C-CON versus WBGT 31°C-FAN, with multiplicity adjusted using the Holm method for p-values and the Bonferroni method for 95% CIs. Within-trial comparisons evaluated pairwise differences between thermoneutral rest and subsequent time points within each trial; these exploratory comparisons were conducted without multiplicity adjustment for p-values or 95% CIs. The level of significance was set at *p* < 0.05. All statistical analyses were performed using R, version 4.6.1 (R Foundation for Statistical Computing, Vienna, Austria).

## 3. Results

Descriptive statistics for all outcomes are provided in the supplementary materials (Supplementary Table S1 and Table S2).

### 3.1 Core Body Temperature

There were no significant differences in rectal temperature among the trials during thermoneutral rest (all *p* = 1.000).

In between-trial comparisons, WBGT 31°C-FAN significantly suppressed the increase in rectal temperature compared with WBGT 31°C-CON, with differences of 0.22°C (95% CI: 0.07 to 0.37°C, *p* = 0.002) at 40 min of exercise and 0.50°C (95% CI: 0.35 to 0.65°C, *p* < 0.001) at 60 min of exercise. Under the WBGT 25°C condition, no significant differences were observed between trials at any time point (difference at 60 min: 0.09°C, 95% CI: −0.06 to 0.24°C, *p* = 0.276).

Similarly, there was no significant difference between WBGT 25°C-CON and WBGT 31°C-FAN (difference at 60 min: 0.05°C, 95% CI: −0.10 to 0.20°C, *p* = 0.431).

In within-trial comparisons, WBGT 31°C-CON showed significant increases of +0.26°C (95% CI: 0.14 to 0.38°C, *p* < 0.001) at 40 min and +0.62°C (95% CI: 0.50 to 0.74°C, *p* < 0.001) at 60 min of exercise. WBGT 25°C-CON also exhibited a significant increase of +0.18°C (95% CI: 0.06 to 0.30°C, *p* = 0.005) at 60 min of exercise. In contrast, in the FAN trials, the increase in rectal temperature throughout the 60-min exercise in WBGT 31°C was limited to +0.11°C (95% CI: −0.01 to 0.23°C), which was not significant (*p* = 0.077). The increase in WBGT 25°C was also non-significant (+0.09°C, 95% CI: −0.03 to 0.21°C, *p* = 0.151) (Figure 2).

**Figure 2.**
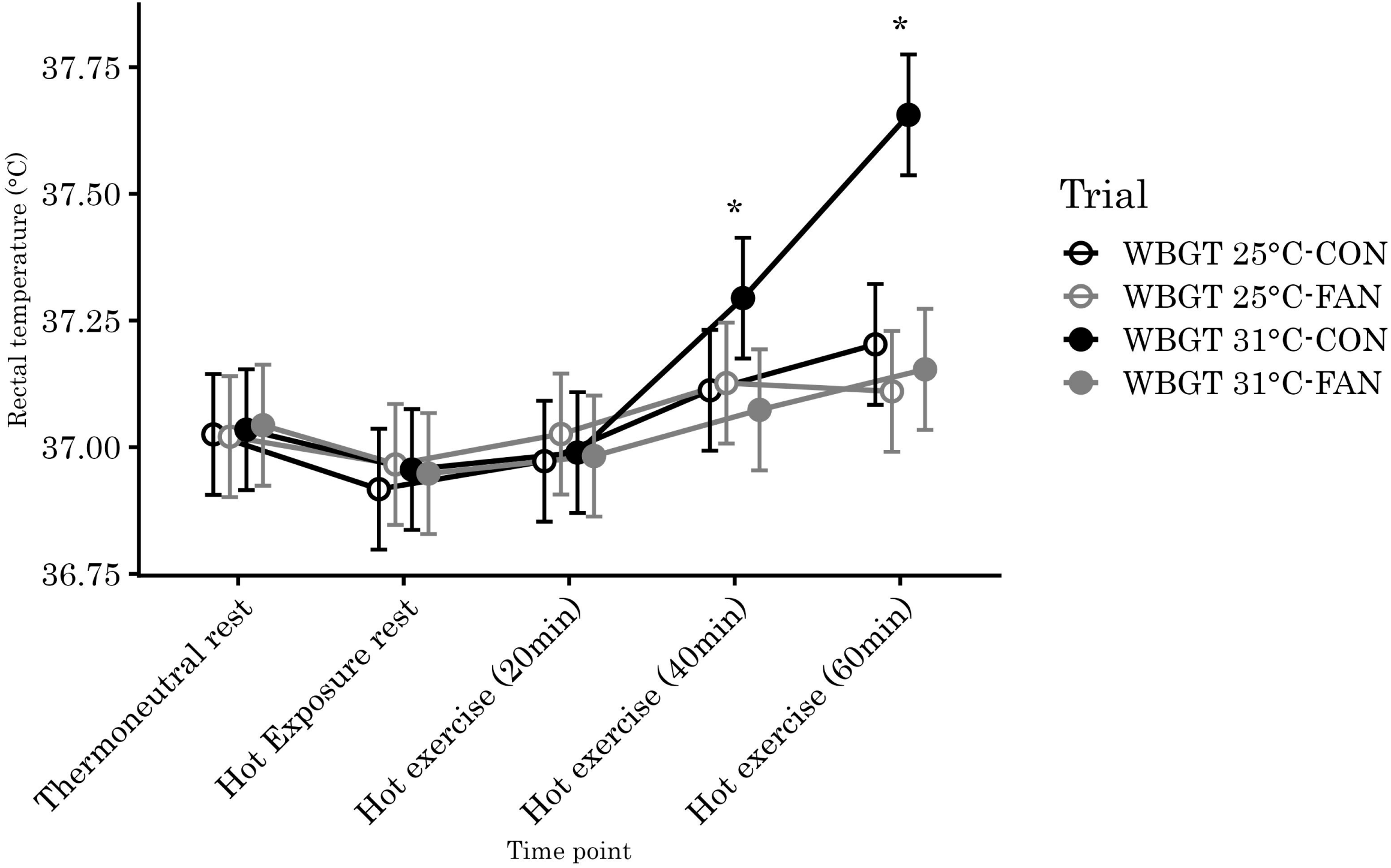
Changes in rectal temperature during thermoneutral rest, hot rest, and 60 min of exercise across the four experimental trials. Values represent estimated marginal means with 95% confidence intervals. * p < 0.05 for WBGT 31°C-CON vs. WBGT 31°C-FAN. CON, control; FAN, fan-attached garment; WBGT, wet bulb globe temperature.

### 3.2 Skin Temperature

There were no significant differences in skin temperatures at any measurement site among the trials during thermoneutral rest (all *p* = 1.000).

For chest skin temperature, between-trial comparisons revealed that WBGT 31°C-FAN was significantly lower than WBGT 31°C-CON by 1.19°C (95% CI: 0.21 to 2.17°C, *p* = 0.008) at 40 min and 1.56°C (95% CI: 0.58 to 2.54°C, *p* < 0.001) at 60 min of exercise. Similarly, in WBGT 25°C, the FAN trial was significantly lower than the CON trial by 1.60°C (95% CI: 0.62 to 2.58°C, *p* < 0.001) at 40 min and 2.55°C (95% CI: 1.57 to 3.53°C, *p* < 0.001) at 60 min of exercise. No significant difference was observed between WBGT 31°C-FAN and WBGT 25°C-CON at 60 min of exercise. Within-trial comparisons showed that chest skin temperature increased significantly from hot rest onward across all trials (all *p* ≤ 0.025). However, in WBGT 25°C-FAN, it returned to the thermoneutral baseline level at 60 min of exercise.

For upper arm skin temperature, WBGT 31°C-FAN was significantly lower than WBGT 31°C-CON by 0.60°C (95% CI: 0.13 to 1.06°C, *p* = 0.008) at 20 min, 0.83°C (95% CI: 0.37 to 1.30°C, *p* < 0.001) at 40 min, and 1.02°C (95% CI: 0.56 to 1.49°C, *p* < 0.001) at 60 min of exercise. In WBGT 25°C, the FAN trial was also significantly lower by 0.48°C (95% CI: 0.02 to 0.94°C, *p* = 0.026) at 40 min and 0.66°C (95% CI: 0.20 to 1.12°C, *p* = 0.002) at 60 min of exercise. No significant differences were observed between WBGT 31°C-FAN and WBGT 25°C-CON. Within-trial comparisons showed a significant increase at all measurement points from hot rest to the end of 60-min exercise across all trials (all *p* < 0.001).

For thigh skin temperature, no significant differences were observed between trials at any time point. Within-trial comparisons indicated that thigh skin temperature increased, remaining significantly elevated from hot rest through the end of 60-min exercise across all trials (all *p* < 0.001).

### 3.3 Heart Rate

There were no significant differences in heart rate among the trials during thermoneutral rest. In between-trial comparisons, WBGT 31°C-FAN resulted in significantly lower heart rates than WBGT 31°C-CON by 15.5 bpm (95% CI: 8.4 to 22.6 bpm, *p* < 0.001) at 20 min, 23.6 bpm (95% CI: 16.5 to 30.7 bpm, *p* < 0.001) at 40 min, and 31.5 bpm (95% CI: 24.4 to 38.6 bpm, *p* < 0.001) at 60 min of exercise. Similarly, in WBGT 25°C, the FAN trial was significantly lower by 10.1 bpm (95% CI: 2.9 to 17.2 bpm, *p* = 0.002) at 40 min and 14.9 bpm (95% CI: 7.7 to 22.1 bpm, *p* < 0.001) at 60 min of exercise. Furthermore, the heart rate in WBGT 31°C-FAN at 60 min of exercise was significantly lower than that in WBGT 25°C-CON by 6.0 bpm (95% CI: −1.2 to 13.1 bpm, *p* = 0.044).

In within-trial comparisons, WBGT 31°C-CON showed a significant increase starting from hot rest (+6.6 bpm, 95% CI: 0.9 to 12.4 bpm, *p* = 0.025), reaching a maximum increase of +68.3 bpm (95% CI: 62.5 to 74.1 bpm, *p* < 0.001) at 60 min of exercise. In contrast, no significant increases were observed during hot rest in WBGT 31°C-FAN, WBGT 25°C-CON, and WBGT 25°C-FAN (*p* = 0.077, *p* = 0.080, and *p* = 0.469, respectively), although all showed significant increases from 20 min of exercise (all *p* < 0.001) (Figure 3).

**Figure 3.**
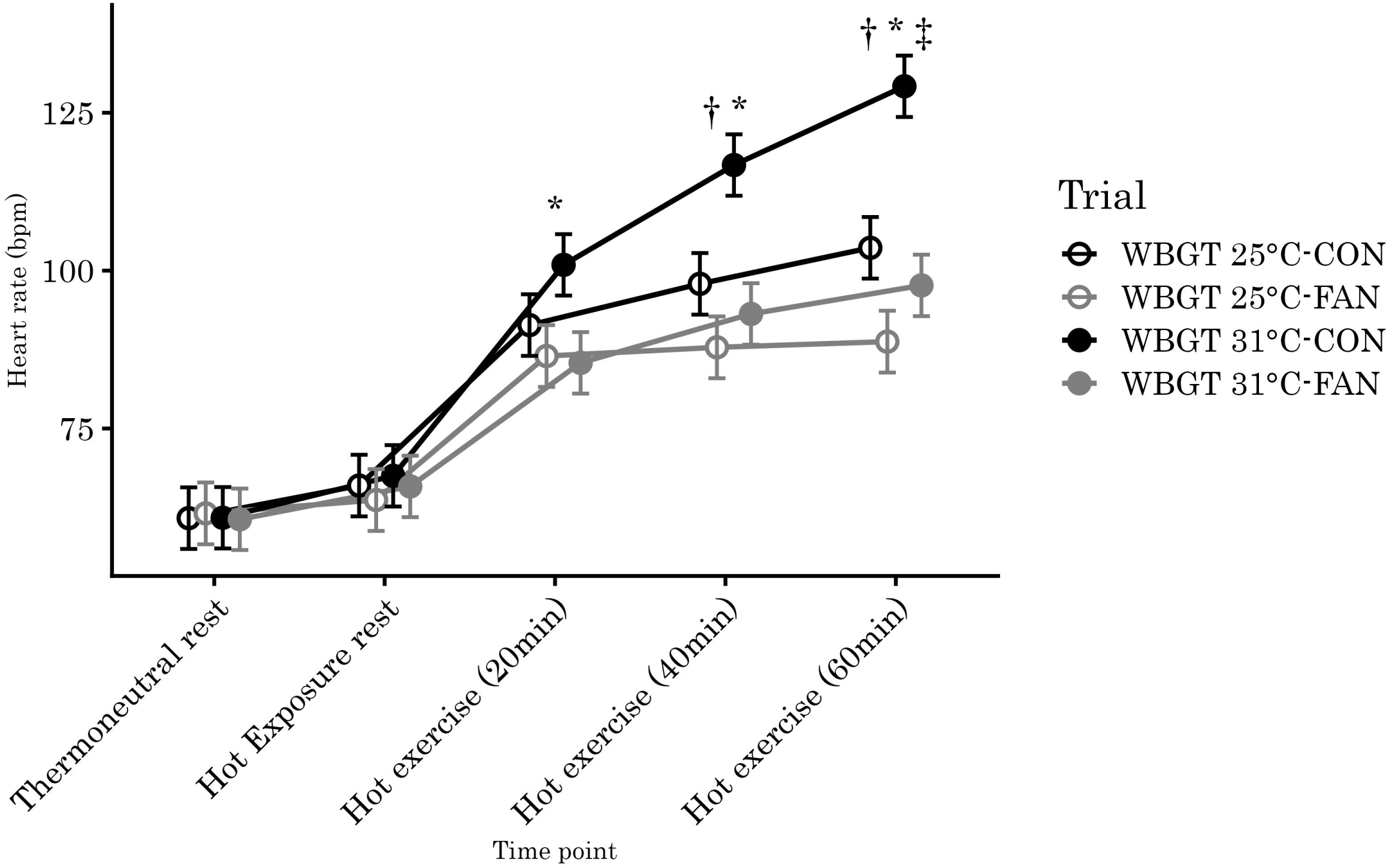
Changes in heart rate during thermoneutral rest, hot rest, and 60 min of exercise across the four experimental trials. Values represent estimated marginal means with 95% confidence intervals. * p < 0.05 for WBGT 31°C-CON vs. WBGT 31°C-FAN; † p < 0.05 for WBGT 25°C-CON vs. WBGT 25°C-FAN; ‡ p < 0.05 for WBGT 25°C-CON vs. WBGT 31°C-FAN. CON, control; FAN, fan-attached garment; WBGT, wet bulb globe temperature.

### 3.4 Microclimate Environment

Microclimate temperature did not differ significantly among the trials during thermoneutral rest (all *p* ≥ 0.733). In between-trial comparisons, no significant differences were observed between WBGT 31°C-FAN and WBGT 31°C-CON. In WBGT 25°C, although the CON trial was significantly lower than the FAN trial (difference: −1.3°C, 95% CI: −2.3 to −0.2°C, p = 0.008) during hot rest due to the introduction of ambient air, no significant differences were observed between trials during exercise. Additionally, from hot rest through all exercise time points, the temperature in WBGT 25°C-CON was significantly lower than in WBGT 31°C-FAN (hot rest: difference: −1.6°C, 95% CI: −2.7 to −0.5°C, p = 0.002; 60 min: difference: −1.2°C, 95% CI: −2.3 to 0.0°C, p = 0.038). Within-trial comparisons showed that microclimate temperature increased significantly from hot rest through the end of 60-min exercise across all trials (all *p* < 0.001).

There were no significant differences in microclimate humidity among the trials during thermoneutral rest (all *p* ≥ 0.598). In between-trial comparisons, WBGT 31°C-FAN was significantly lower than WBGT 31°C-CON at all time points from hot rest (difference: 13.7%, 95% CI: 5.8 to 21.7%, *p* < 0.001) to the end of 60-min exercise (difference: 18.8%, 95% CI: 10.5 to 27.2%, *p* < 0.001). Similarly, in WBGT 25°C, the FAN trial was significantly lower than the CON trial at all time points from hot rest (difference: 33.0%, 95% CI: 24.7 to 41.3%, *p* < 0.001) to 60 min of exercise (difference: 39.0%, 95% CI: 30.7 to 47.3%, *p* < 0.001). Furthermore, at all time points during exercise, WBGT 31°C-FAN was significantly lower than WBGT 25°C-CON (20 min: difference: 10.3%, 95% CI: 1.8 to 18.7%, *p* = 0.004; 40 min: difference: 15.1%, 95% CI: 6.6 to 23.5%, *p* < 0.001; 60 min: difference: 17.8%, 95% CI: 9.3 to 26.3%, *p* < 0.001). In within-trial comparisons, both WBGT 31°C-CON and WBGT 25°C-CON showed significant increases from hot rest (WBGT 31°C: +8.2%, 95% CI: 1.8 to 14.6%, *p* = 0.013; WBGT 25°C: +7.5%, 95% CI: 0.6 to 14.5%, *p* = 0.035), reaching increases of +15.7% (95% CI: 9.0 to 22.4%, *p* < 0.001) and +22.3% (95% CI: 15.3 to 29.2%, *p* < 0.001) at 60 min of exercise, respectively. Conversely, WBGT 25°C-FAN showed a significant decrease relative to thermoneutral rest at all time points from hot rest (−26.7%, 95% CI: −33.1 to −20.2%, *p* < 0.001) through 60 min of exercise (−17.9%, 95% CI: −24.4 to −11.5%, *p* < 0.001). In WBGT 31°C-FAN, no significant changes were observed relative to thermoneutral rest at any time point from hot rest through 60 min of exercise (difference at 60 min: 0.0%, 95% CI: −6.5 to 6.4%, *p* = 0.994) (Figure 4).

**Figure 4.**
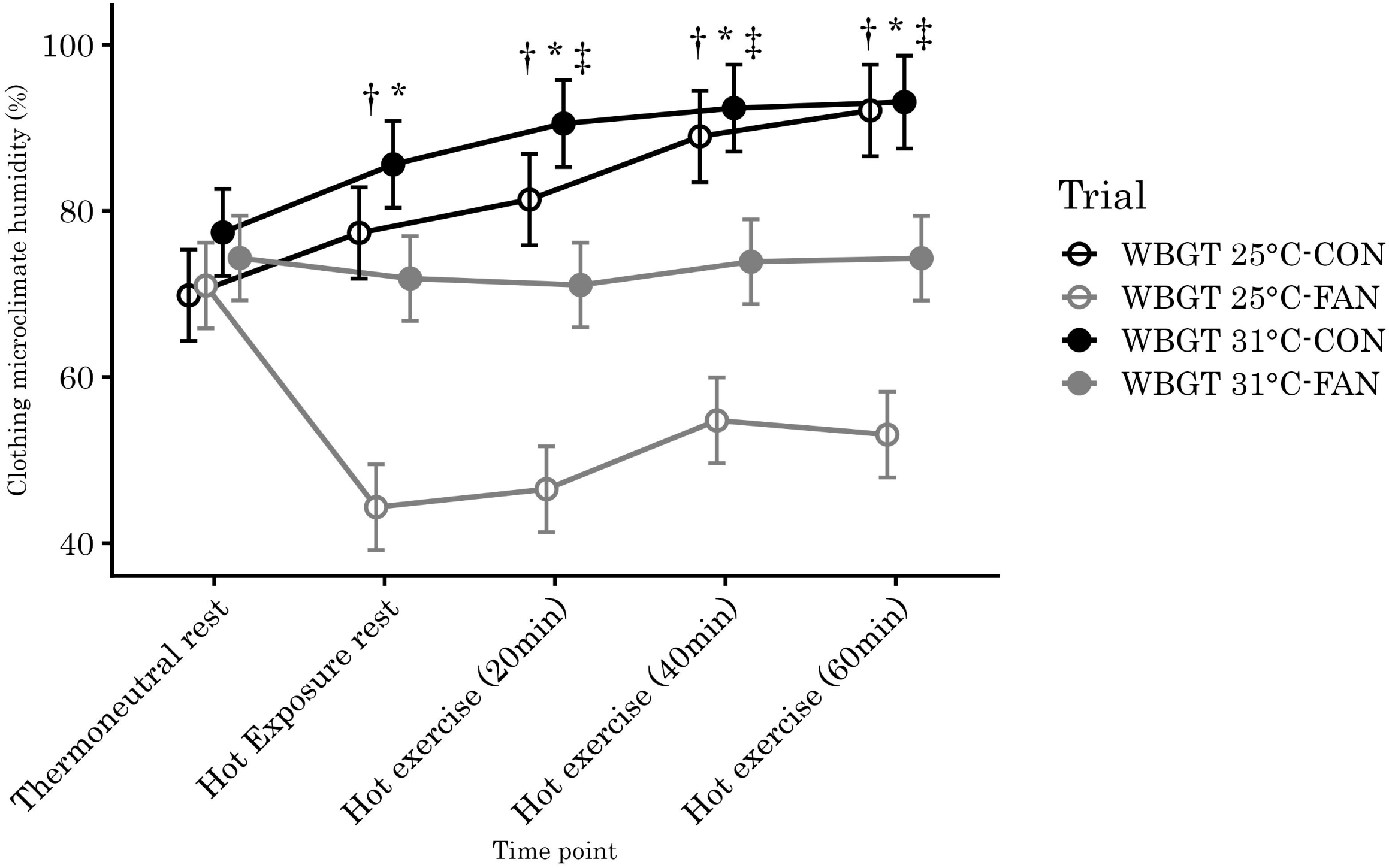
Changes in clothing microclimate humidity during thermoneutral rest, hot rest, and 60 min of exercise across the four experimental trials. Values represent estimated marginal means with 95% confidence intervals. * p < 0.05 for WBGT 31°C-CON vs. WBGT 31°C-FAN; † p < 0.05 for WBGT 25°C-CON vs. WBGT 25°C-FAN; ‡ p < 0.05 for WBGT 25°C-CON vs. WBGT 31°C-FAN. CON, control; FAN, fan-attached garment; WBGT, wet bulb globe temperature.

### 3.5 Body Mass Changes

In between-trial comparisons, body mass loss was significantly lower in WBGT 31°C-FAN (0.5 ± 0.2 kg) than in WBGT 31°C-CON (1.0 ± 0.4 kg), with a difference of 0.5 kg (95% CI: 0.2 to 0.7 kg, *p* = 0.001). In WBGT 25°C, no significant difference was observed between CON (0.6 ± 0.2 kg) and FAN (0.4 ± 0.2 kg). There was also no significant difference between WBGT 31°C-FAN and WBGT 25°C-CON.

### 3.6 Subjective Outcomes

A significant trial-by-time interaction was observed for thermal sensation (*p* = 0.007). However, between-trial comparisons revealed no significant differences, although the median at 60 min of exercise was 9 (very hot) in WBGT 31°C-CON and 8 in WBGT 31°C-FAN (*p* = 0.093). In within-trial comparisons, significant increases were observed from hot rest in WBGT 25°C-CON (*p* = 0.021), WBGT 31°C-CON (*p* = 0.021), and WBGT 31°C-FAN (*p* = 0.048), and from 20 min of exercise in WBGT 25°C-FAN (*p* = 0.034).

No significant trial-by-time interactions were observed for thermal comfort, RPE, and thirst sensation. Furthermore, between-trial comparisons revealed no significant differences at any time point (thermal comfort: *p* ≥ 0.053, RPE: *p* ≥ 0.102, thirst sensation: *p* ≥ 0.259).

## 4. Discussion

The results showed that the fan-attached garment significantly suppressed the elevation of core body temperature during exercise in a humid environment. Furthermore, microclimate humidity decreased under low humidity and did not increase during exercise even under high humidity.

These findings suggest that high-airflow fan-attached garments reduce physiological strain during physical activity in hot environments.

Notably, the FAN trial almost completely suppressed core body temperature increases in a hot and humid environment of WBGT 31°C. Excessive increases should be assessed against international occupational safety and health standards. ISO 7243 establishes WBGT reference values according to metabolic rate, based on a maximum permissible rectal temperature of 38.0°C for healthy workers^16^. Given that the resting core body temperature of healthy adults is approximately 37.0°C, the allowable increase to avoid heat-related illness or excessive heat stress is approximately 1.0°C. Previous studies of fan-attached garments primarily compared them with control conditions and did not necessarily assess within-trial changes; however, their reported values show an increase in rectal temperature of >0.5°C per hour in the fan trials, even under exercise intensities and environmental conditions comparable to those of the present study^5,6^. This level of heat accumulation could reach the safety limit over the course of a working day, even with rest breaks. Conversely, in the FAN trial, no significant increase in rectal temperature was observed throughout the 1-h continuous exercise. In the between-trial comparison at WBGT 31°C, a significant suppression of 0.5°C in estimated marginal means was confirmed. This pronounced suppression was likely attributable to the airflow rate used in this study (80 L/s), which was substantially higher than those examined in previous studies (approximately 12–30 L/s) and provided marked forced convection and ventilation capacity.

Marked improvements were also observed in cardiovascular strain and sweat response. At WBGT 31°C, FAN reduced heart rate during exercise by up to 32 bpm and decreased body mass loss by 0.5 kg compared with CON. This suggests that the enhancement of evaporative heat dissipation by high airflow alleviates cardiovascular strain and dehydration risk. Toward the end of the exercise, the heart rate in WBGT 31°C-FAN was 6 bpm lower than that in WBGT 25°C-CON, an environment with lower heat stress. This suggests that wearing a high-airflow fan-attached garment in a humid and hot environment may reduce physiological strain more effectively than performing physical activity without a fan in a low-humidity hot environment. A difference in microclimate humidity of approximately 10% has been proposed to have no substantial effect on systemic physiological responses^7,8^. In the present study, wearing the fan-attached garment substantially reduced humidity under low-humidity conditions and maintained microclimate humidity at low levels under high-humidity conditions, indicating that the high airflow of 80 L/s effectively expelled sweat vapor from the garment and promoted evaporative cooling. Thus, preventing the exercise-induced increase in microclimate humidity under hot and humid conditions likely mediated the suppression of core temperature elevation and physiological strain.

This study has some limitations. First, the participants were exclusively healthy men aged 18–45 years. The generalizability of these findings to women and older workers, whose sweating and skin blood flow responses during heat stress are diminished with advancing age^17^, warrants further investigation. Second, this study was conducted in a controlled laboratory environment rather than in an actual field setting, which limits its external validity. However, the exercise workload corresponds to approximately 5 metabolic equivalents (METs)^18^, which is comparable to moderate-to-vigorous physical tasks in industrial settings such as construction and manufacturing. Therefore, our findings provide empirical evidence under conditions that adequately simulated metabolic heat production and physiological strain encountered in demanding work environments. Third, the sample size was small, and no conventional-airflow condition was included; therefore, comparisons with lower airflow rates rely on previous studies. Field studies in outdoor and industrial workplaces and in diverse populations are needed to confirm these findings.

These findings have important implications for occupational health management. In Japan, the revised Ordinance on Industrial Safety and Health, enforced in 2025, requires employers to establish and disseminate reporting systems and procedures for preventing severe heat-related illnesses for work anticipated to last continuously for ≥1 h (or >4 h per day) in environments with WBGT ≥28°C or ambient temperature ≥31°C^19^. The present study demonstrates that a high-airflow fan-attached garment can substantially suppress core body temperature elevation in a demanding 1-h continuous heat-stress work model corresponding to these regulatory criteria. Additionally, the garment operates continuously for 4 h at the maximum airflow of 80 L/s (manufacturer-reported specification); with a spare battery after the morning shift, it can therefore be used at maximum airflow throughout the workday. Therefore, where fan-attached garments are permitted, high-airflow models may serve as a practical occupational health tool to maintain workers’ health and productivity in hot conditions.

## Data Availability

The data underlying this article are available in the article and in its online supplementary material.

## Acknowledgments

We express our sincere gratitude to all the participants who cooperated with the protocol of this study.

## Author Contributions

F.T. conceived the study and administered the project; K.H. designed the methodology; K.H., M.W., and Y.U. collected the data; M.W. curated the data; K.H. and H.I. analyzed the data and created the visualizations; Y.T. supervised the study; K.H. and F.T. drafted the original manuscript; and M.W., H.I., Y.U., and Y.T. reviewed and edited the manuscript.

