## Supplementary material for "Effects of a High-Airflow Fan-Attached Garment on Thermoregulatory and Physiological Responses During Physical Activity in Hot Environments: A Randomized Crossover Trial": Figure S1

**Enrolment**

**Analysis**

**Follow-Up**

Analyzed for primary outcome (n = 7)

Excluded from analysis (n = 0)

Discontinued intervention (n = 0)

Lost to follow-up for primary outcome (n = 0)

**Allocation**

Excluded (n = 1)

Not meeting inclusion criteria (n = 1)

Declined to participate (n = 0)

Other reasons (n = 0)

Randomized (n = 7)

Assessed for eligibility (n = 8)

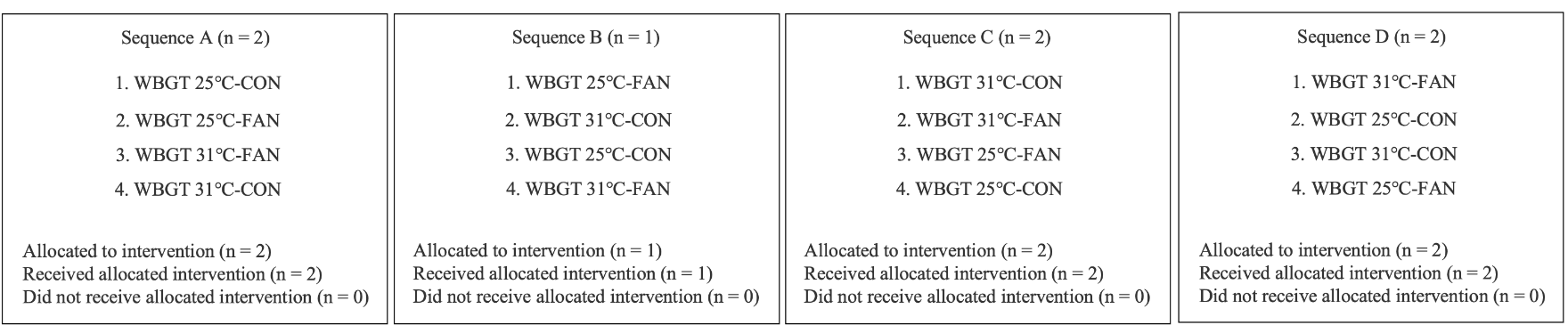

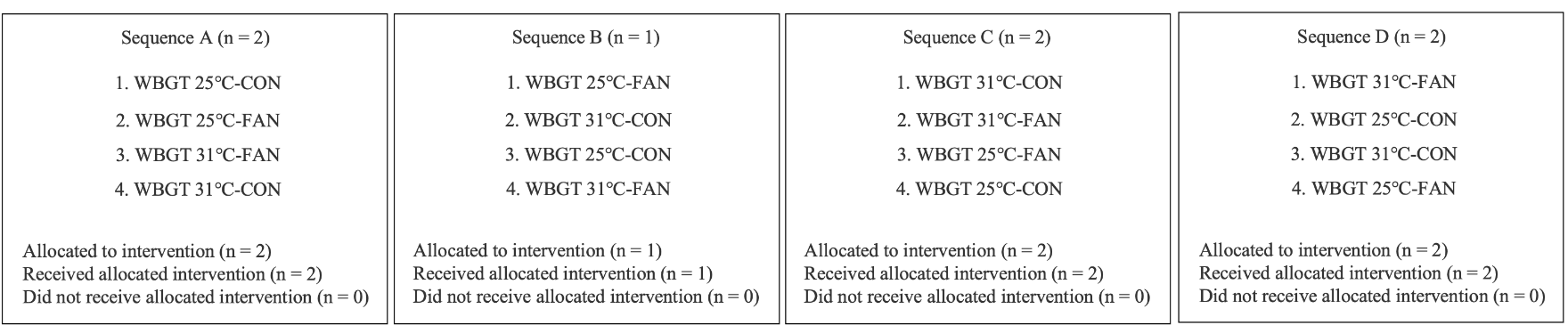

**Supplementary Figure S1.** CONSORT flow diagram of the crossover trial. Participants were randomized into four sequences. CON, control garment; FAN, fan-attached garment; WBGT, wet bulb globe temperature.
